# Categorical and dimensional brain network-based models of trauma-related dissociative subtypes

**DOI:** 10.1101/2022.04.29.22274474

**Authors:** Lauren A. M. Lebois, Poornima Kumar, Cori A. Palermo, Ashley M. Lambros, Lauren O’Connor, Jonathan D. Wolff, Justin T. Baker, Staci A. Gruber, Nina Lewis-Schroeder, Kerry J. Ressler, Matthew A. Robinson, Sherry Winternitz, Lisa D. Nickerson, Milissa L. Kaufman

**Author notes:** **Address correspondence to:** Lauren A. M. Lebois, Ph.D., Division of Depression and Anxiety, McLean Hospital / Harvard Medical School, 115 Mill St, Belmont MA 02478. co-first (Lauren A. M. Lebois, Poornima Kumar). co-last (Lisa D. Nickerson, Milissa L. Kaufman).

## Abstract

**Background:** Trauma-related pathological dissociation is a multidimensional and disabling phenomenon that involves disruptions or discontinuities in psychological functioning. Despite its prevalence, personal and societal burden, dissociation remains underappreciated in clinical practice, and it lacks a synthesized neurobiological model that could place it in context with other common psychiatric symptoms. To identify a nuanced neurobiological model of pathological dissociation, we examined the functional connectivity of three core neurocognitive networks as related to the dimensional dissociation subtypes of depersonalization/derealization and partially-dissociated intrusions, and the diagnostic category of a complex dissociation disorder, dissociative identity disorder (DID).

**Methods:** Participants were 91 adult women with and without: a history of childhood trauma, current posttraumatic stress disorder (PTSD) and varied levels of pathological dissociation. Participants provided interview and self-report data about pathological dissociation, PTSD symptoms, childhood maltreatment history, and completed a resting-state functional magnetic resonance imaging scan.

**Results:** After controlling for age, childhood maltreatment and PTSD symptom severity, we found that pathological dissociation was associated with hyperconnectivity within central executive, default, and salience networks, and decreased connectivity of central executive and salience networks with other areas. Moreover, we isolated unique connectivity markers linked to depersonalization/derealization, to partially-dissociated intrusions, and to DID.

**Conclusions:** Our work suggests subtypes of pathological dissociation have robust, discernable, and unique functional connectivity signatures. The neural correlates of dissociation may serve as potential targets for treatment engagement to facilitate recovery from PTSD and pathological dissociation. These results underscore dissociation assessment as crucial in clinical and medical care settings.

## Introduction

Pathological dissociation is a common experience in the aftermath of trauma (1). Namely, it is a central feature of both the dissociative subtype of posttraumatic stress disorder (PTSD) and dissociative identity disorder (DID) (2). These conditions are surprisingly prevalent: PTSD has a lifetime prevalence upwards of 12% and 15-30% of those people will have the dissociative subtype (3). Similarly, up to 3% of the population will suffer from DID in their lifetime (4). Despite their prevalence, these conditions remain understudied and often misunderstood.

Research embracing trauma-related pathological dissociation has determined that it encompasses a range of disruptions or discontinuities in someone’s psychological experience (2). General symptoms like depersonalization and derealization are frequent experiences in both the dissociative subtype of PTSD and DID (2). Depersonalization and derealization involve feelings of detachment or disconnection from one’s sense of self, body and environment (2). When experiencing depersonalization, individuals report feeling like their body is unreal or feels like it is not their own. Likewise, when experiencing derealization, individuals report feeling that their environment is unreal or like they are in a movie. Dissociation also includes experiences of self-alteration, for example, partially-dissociated intrusions, that are common in DID (5). When experiencing partially-dissociated intrusions, individuals report feeling like they are hearing voices or that their thoughts, emotions, and actions emerge without their control. These intrusions are “partially-dissociated “ because the person is aware of these experiences, but they feel like the experiences do not belong to them (5).

Both general dissociative symptoms and experiences of self-alteration can help people cope in the face of inescapable threat and trauma (6); however, they can also impede one’s ability to function and can interfere with new emotional learning (7). Effective treatments exist; yet symptoms of dissociation and dissociative disorders frequently go undiagnosed or misdiagnosed (8).

Brain-based measures of dissociation can provide scientific evidence for the validity of these experiences and can link the clinical phenomenology with biological mechanisms. While foundational studies have begun to characterize the neurobiology of dissociation (3), the field lacks a synthesized model that could place it in context with other common psychiatric conditions.

### The Triple Network Model of Psychopathology

The Triple Network model of psychopathology may be able to provide a synthesized neurobiological model for pathological dissociation. This model offers an integrative framework based in systems neuroscience for understanding cognitive and affective dysfunction across psychiatric conditions (9). The basic model implicates altered intrinsic organization and interactions between three large-scale brain networks across disorders: the central executive (CEN), default (DN), and salience networks (SN).

These three networks serve complementary functions. The CEN, a lateral frontoparietal network, is involved in tasks that are cognitively challenging, for example, working memory, problem solving, and goal-directed decision making (10). Conversely, DN, a medial frontoparietal network (10), is involved in self-generated thought, for example, past and future thinking, self-referential thinking, autobiographical memory, and daydreaming (11,12). Lastly, SN, a midcingulo-insular network (10), is involved in perceiving relevant autonomic and emotional information (9). Consequently, the SN initiates shifts between CEN and DN – helping to determine one’s focus of attention (13).

There is also convergent evidence that CEN, DN, and SN are heterogeneous systems comprised of subnetworks. In the Triple Network model, the sub-systems represented are: 1) The medial temporal subnetwork of the DN (tDN; (11,14–16)). Among other regions, tDN includes hippocampus and amygdala and is engaged during autobiographical memory, recollection of events in one’s past (i.e., episodic memory retrieval) and simulating future events. 2) The cingulo-opercular subnetwork of the SN (cSN). This subnetwork is engaged during interoception, especially the experience of emotion derived from information about the internal milieu (13). It is also a transitional network, linking cognition, emotion, and interoception (17,18). 3) The right-lateralized CEN (rCEN). This subnetwork is strongly implicated in cognitive processes such as reasoning, attention, inhibition, and memory (17,19). The rCEN is distinct from left CEN, which is primarily involved in language processing (17,19). These subnetworks are easily identified using group independent component analysis and are highly reproducible (17,19,20).

### Functional Connectivity of Trauma-related Pathological Dissociation

Altered organization and interaction between CEN, DN, and SN are consistently reported across psychiatric disorders (9). Central to these alterations is improper assignment of relevance or salience to either internal or external stimuli (9). Inappropriate salience detection, failing to assign relevance to something important or assigning relevance to something unnecessarily, can create a cascade effect where the CEN and DN do not engage or disengage appropriately. Depending on the subtype of pathological dissociation, these symptoms could involve inappropriate salience detection in either direction, and concomitant alterations in executive functioning and self-generated thought.

Although the Triple Network model of psychopathology has not yet been applied to dissociation, neuroimaging work to date implicates altered connectivity of regions in all three networks (21,22). These studies typically focus on more general dissociative symptoms of depersonalization and derealization – with seed-based functional connectivity findings in the dissociative subtype of PTSD highlighting altered connectivity of regions located in the SN, DN, and CEN (e.g., amygdala, insula, prefrontal and parietal cortex (3)). One study from our team found that hyperconnectivity of regions in CEN and DN was associated with a measure of pathological dissociation that combined scores of depersonalization, derealization, and partially-dissociated intrusions in a PTSD, PTSD dissociative subtype, and DID sample (23). Taken together, these findings cover a range of dissociation subtypes; however, they do not directly compare different subtypes. The unique contributions of different dissociation subtypes to altered connectivity in the three core networks of the Triple Network model are unknown.

### Experiment Overview

To address this gap, we assessed the connectivity of rCEN, tDN, and cSN (**Fig. S1**) as related to different subtypes of pathological dissociation: the dimensional symptoms of depersonalization/derealization and partially-dissociated intrusions, and the diagnostic category of DID using a novel method for assessing both overlapping and unique contributions of different dissociation types (24). Given prior work and the Triple Network model of psychopathology, we hypothesized all three networks would be implicated in dissociation and unique patterns of connectivity would emerge for each dissociative subtype.

## Methods and Materials

### Participants

A total of 109 adult women with and without histories of childhood trauma, current PTSD, and varied levels of dissociative symptoms were enrolled. Of these, a total of 18 datasets were excluded from analysis due to data quality issues, resulting in 91 datasets available for subsequent analysis. See **Table 1** demographic and clinical measures. Participants with PTSD (*N*=65) had histories of childhood trauma and varied levels of pathological dissociation, including some with the PTSD dissociative subtype, and some with DID. These individuals were seeking treatment at a psychiatric hospital in the northeast region of the US. Participants without PTSD had no history of or current psychiatric disorders (*N*=26).

**Table 1.** Demographics and clinical characteristics.

|  | Control | Conventional PTSD | PTSD Dissociative Subtype | DID* |
| --- | --- | --- | --- | --- |
|  | *N* | *N* | *N* | *N* |
| Sample Size | 28 | 19 | 18 | 26 |
| Race |  |  |  |  |
| American Indian | 0 | 0 | 0 | 1 |
| Asian | 1 | 0 | 1 | 3 |
| Black / African American | 2 | 1 | 0 | 2 |
| White | 25 | 18 | 16 | 20 |
| Other | 0 | 0 | 1 | 0 |
| Ethnicity |  |  |  |  |
| Hispanic / Latinx | 3 | 1 | 0 | 1 |
| Non-Hispanic / Latinx | 25 | 18 | 17 | 25 |
| Prefer not to answer | 0 | 0 | 1 | 0 |
| Education |  |  |  |  |
| Grade 7 to 12 (without graduating high school) | 0 | 0 | 0 | 1 |
| High school or equivalent | 1 | 0 | 1 | 0 |
| Part of College | 3 | 9 | 4 | 9 |
| College (2 year) | 0 | 0 | 2 | 1 |
| College (4 year) | 12 | 3 | 5 | 5 |
| Part of Graduate/Professional School | 4 | 3 | 4 | 3 |
| Completed Graduate/Professional School | 8 | 4 | 2 | 7 |
|  | *M ± SD* | *M ± SD* | *M ± SD* | *M ± SD* |
| Age | 32.2 ± 11.2 | 33.4 ± 10.9 | 29.5 ± 9.6 | 37.4 ± 13.6 |
| CTQ Total Severity | 29.5 ± 6.6 | 63.6 ± 20.5 | 75.4 ± 22.6 | 88.1 ± 13.5 |
| CAPS-5 Total Severity | 0.64 ± 1.5 | 48.1 ± 10.6 | 48.5 ± 9.9 | 52.9 ± 13.6 |
| PCL-5 Total | 2.0 ± 4.4 | 53.9 ± 10.5 | 53.2 ± 12.3 | 50.5 ± 16.0 |
| MID Severe Pathological Dissociation | 2.2 ± 1.8 | 44.2 ± 22.6 | 76.3 ± 24.8 | 117.9 ± 29.2 |
| MID Depersonalization/Derealization | 0.51 ± 0.96 | 12.4 ± 9.2 | 30.1 ± 15.1 | 42.9 ± 19.6 |
| MID Partially-Dissociated Intrusions | 0.57 ± 0.66 | 13.9 ± 8.7 | 20.9 ± 12.9 | 45.2 ± 16.2 |

**Note.** \*All participants with DID also met criteria for the dissociative subtype of PTSD. Total sample *N* = 91. PTSD, posttraumatic stress disorder; CTQ, childhood trauma questionnaire; CAPS-5, Clinician-Administered PTSD Scale for DSM 5; PCL-5, PTSD Checklist for DSM 5; MID, multidimensional inventory of dissociation

All research procedures were approved by the Massachusetts General Brigham Human Research Affairs Institutional Review Board and performed in accordance with human subject guidelines and regulations. All participants provided written informed consent and received $200 compensation.

### Diagnostic and Symptom Measures

Data collection followed the STROBE guidelines (25). Psychiatric diagnoses were determined using the Structured Clinical Interview for the DSM-IV Axis I Disorders (26), the Clinician-Administered PTSD Scale for DSM-5 (CAPS-5; (27)) and the Structured Clinical Interview for DSM-IV Dissociative Disorders-revised (28).

For a measure of general dissociative symptoms, we used the average of the depersonalization and derealization subscales on the Multidimensional Inventory of Dissociation (MID; (29)). To measure partially-dissociated intrusions, we used the average of several MID subscales (see supplement). To control for PTSD symptom and childhood maltreatment severity in our analyses, we used the CAPS-5 total PTSD symptom severity score and the Childhood Trauma Questionnaire (CTQ) total score (30).

### MRI Procedures

See the supplement for detailed information on MRI procedures, data quality assurance, preprocessing, and statistical analysis. We acquired resting-state fMRI data and conducted standardized preprocessing using *fMRIPrep* 20.0.1 (24,31). Resting-state networks were derived using group Independent Component Analysis (GICA; (32)). We then implemented a dual regression approach to obtain subject-specific network maps corresponding to each GICA component (33). We identified rCEN, cSN, and tDN by visual inspection of the spatial maps to find the components with the greatest spatial overlap with previously reported networks (17,19,34) (**Fig. S1**).

### Categorical and Dimensional Connectivity Analysis

DID diagnosis and the dimensional symptoms of depersonalization/derealization and partially-dissociated intrusions are highly collinear. Evaluating associations between each dissociation type and network connectivity in separate models could yield findings that are driven by shared variance due to their collinearity (**Fig. 1**). On the other hand, evaluating all three predictors within the same model will reduce sensitivity because shared variance between the predictors is ignored in the estimation.

**Figure 1.**
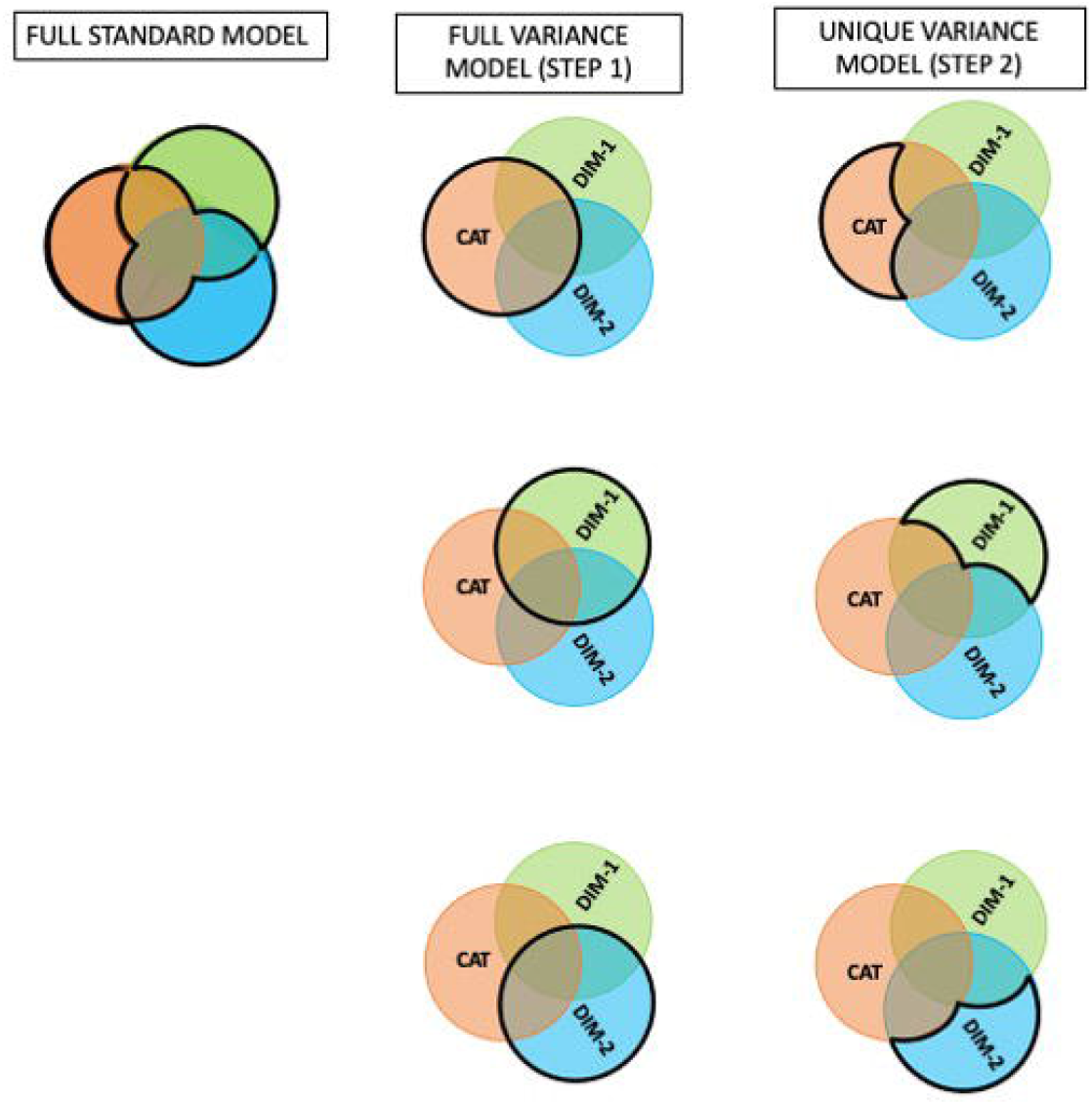
Statistical Approach of the Full and Unique Variance models. Full standard model represents a multiple regression model that includes all diagnostic categorical and dimensional variables together. In this case, the shared variance between the variables (the areas of overlap in the center of the Venn diagram) are ignored when estimating the regression parameters. In contrast, full variance modeling (Step 1) involves running separate models that estimate the regression parameter of the variable using its full variance (heavy black circles) to yield a set of brain regions (or markers) whose connectivity with the network is associated with that variable. In Step 2, unique variance modeling identifies the unique association between the markers identified in Step 1 and each diagnostic categorical and dimensional variable. Adapted from (24).

To address these issues, we followed the novel two-step approach presented in (24) that relied on a series of multiple regressions with orthogonalization of predictors. First, full variance models were estimated with each network’s set of connectivity maps as the dependent variable, and orthogonalized predictors of interest (i.e., DID, depersonalization/derealization, and partially-dissociated intrusions). This identified brain regions or markers associated with each variable using the full variance associated with the variable. We interpret these markers as being associated with “pathological dissociation,” irrespective of subtype.

We then identified unique contributions of each dissociation subtype to the connectivity between markers and the network(s) by extracting the subject-level average regression weights for each marker. These weights were then used as dependent variables in a second set of multiple regressions with orthogonalizations to estimate regression coefficients that captured the unique effects of each subtype (see **Fig. 1** and Supplement for full details).

Each full variance model had one of the following independent variables as the predictor: diagnostic subgroup (PTSD, PTSD dissociative subtype, DID, and control) and two additional symptom severity scores (depersonalization/derealization, partially-dissociated intrusions). In addition, age, CTQ childhood maltreatment severity, and CAPS-5 PTSD total symptom severity were entered as covariates of no interest in all models. Every full variance model was evaluated using FSL Randomize for non-parametric permutation testing (*n*=5000 permutations) with threshold-free cluster enhancement to control family-wise error (*p*<.05). As noted in (24), an additional correction for the multiple regression models is not necessary because all models explain the same total variance and, as such, are equivalent with respect to considerations of signal versus noise. Further, as in (24), unique variance models were not corrected to retain sensitivity.

## Results

### Right Central Executive Network (rCEN)

Full variance models showed the rCEN was most impacted by pathological dissociation; specifically, 39 clusters were linked to two types of alterations: 1) within-network hyperconnectivity; 2) Decreased connectivity with brain regions outside rCEN (**Fig. 2, Table 2**).

**Table 2.** Connectivity in Default, Salience and Central Executive Network related to Pathological Dissociation

| Cluster Index | Brain Region | Network of Brain Region | Direction of Full Variance Relationship | Full Variance Model | Cluster size | MNI coordinates (x, y, z) | Unique variance contributions |  |  |
| --- | --- | --- | --- | --- | --- | --- | --- | --- | --- |
|  |  |  |  |  |  |  | DID Diagnosis t (p)<br>N=54 | Depersonalization/<br>Derealization t (p)<br>N=91 | Partially-dissociated Intrusions t (p)<br>N=91 |
| Medial Temporal Default Network |  |  |  |  |  |  |  |  |  |
| 1 | L Parahippocampal gyrus | tDN | DID | DID-HC | 36 | -28, -44, -12 | -1.88 (0.064) | **2.13 (0.036)\*** | -0.45 (0.652) |
| 2 | R Retrosplenial | tDN | DID | DID-HC | 195 | 6, -50, 8 | -0.04 (0.970) | 0.45 (0.656) | -0.20 (0.841) |
| 3 | R Parahippocampal gyrus | tDN | + | Dep/Der | 37 | 28, -40, -14 | -1.57 (0.121) | **2.97 (0.004)\*** | -1.42 (0.163) |
| 4 | L Parahippocampal gyrus | tDN | + | Dep/Der | 90 | -26, -46, -12 | -1.84 (0.070) | **2.10 (0.039)\*** | -0.60 (0.549) |
| 5 | L Retrosplenial, vPCC, Precuneus | tDN | + | Dep/Der | 395 | -14, -62, 16 | 0.67 (0.505) | -0.89 (0.374) | 1.37 (0.175) |
| 6 | R Retrosplenial, vPCC, Precuneus | tDN | + | Dep/Der | 774 | 6, -50, 8 | 0.41 (0.680) | 0.16 (0.875) | 0.32 (0.753) |
| 7 | R middle Occipital gyrus, middle temporal gyrus, Angular Gyrus | tDN | + | PDI | 54 | 48, -64, 245 | -1.30 (0.197) | -0.33 (0.742) | 0.41 (0.680) |
| 8 | L Parahippocampal gyrus, Fusiform gyrus | tDN | + | PDI | 113 | -28, -44, -12 | -1.84 (0.070) | 1.62 (0.109) | -0.33 (0.739) |
| 9 | R Parahippocampal gyrus, Fusiform gyrus | tDN | + | PDI | 172 | 28, -40, -14 | -1.31 (0.192) | 1.94 (0.056) | -0.81 (0.421) |
| 10 | R Retrosplenial, vPCC, Precuneus | tDN | + | PDI | 1802 | 6, -50, 8 | 0.61 (0.543) | -0.69 (0.495) | 1.12 (0.264) |
| Salience Network |  |  |  |  |  |  |  |  |  |
| 11 | L Frontal operculum, insula | cSN | + | Dep/Der | 453 | -32, 12, 4 | 0.46 (0.649) | 0.68 (0.497) | 0.95 (0.345) |
| 12 | R Frontal pole, vIPFC | cSN | + | Dep/Der | 997 | 32, 60, 12 | 0.09 (0.931) | -0.01 (0.995) | 1.34 (0.183) |
| 13 | L Frontal pole, vIPFC, dIPFC, supplementary motor area, dACC<br>R dIPFC, supplementary motor area, dACC | cSN | + | Dep/Der | 5454 | -34, 44, 10 | 0.66 (0.511) | 0.44 (0.658) | 1.64 (0.105) |
| 14 | R Superior Parietal Lobule | rCEN | - | Dep/Der | 44 | 32, -70, 58 | 1.93 (0.057) | 0.11 (0.914) | -1.03 (0.307) |
| 15 | L Frontal operculum, insula | cSN | + | PDI | 367 | -36, 16, -2 | 0.51 (0.615) | 0.33 (0.744) | 1.29 (0.201) |
| 16 | R Frontal pole, vIPFC | cSN | + | PDI | 1280 | 32, 58, 12 | 0.23 (0.816) | -0.18 (0.859) | 1.47 (0.146) |
| 17 | L Frontal pole, vIPFC, dIPFC, supplementary motor area, dACC<br>R supplementary motor area, dACC | cSN | + | PDI | 5852 | -20, 52, 14 | 0.77 (0.444) | 0.03 (0.974) | 1.95 (0.054) |
| 18 | R Superior Parietal Lobule | rCEN | - | PDI | 54 | 30, -72, 58 | 1.91 (0.059) | 0.18 (0.857) | -1.12 (0.266) |

| Cluster index | Brain Region | Network of Brain Region | Direction of Full Variance Relationship | Full Variance Model | Cluster size | MNI coordinates (x, y, z) | Unique variance contributions |  |  |
| --- | --- | --- | --- | --- | --- | --- | --- | --- | --- |
|  |  |  |  |  |  |  | DID Diagnosis<br>*N*=54 | Depersonalization/<br>Derealization<br>*N*=91 | PDI<br>*N*=91 |
| Right Central Executive Network |  |  |  |  |  |  |  |  |  |
| 19 | L ACC | cSN | HC | HC–DID | 14 | -8, 20, 40 | 1.96 (0.054) | -0.33 (0.743) | 0.07 (0.941) |
| 20 | L Inferior frontal gyrus | --- | HC | HC-DID | 18 | -52, 20, 22 | **2.04 (0.045)\*** | -0.79 (0.432) | 0.87 (0.389) |
| 21 | L Mid cingulate cortex | cSN | HC | HC– DID | 19 | 0, -4, 46 | 1.74 (0.086) | 0.38 (0.705) | 0.12 (0.908) |
| 22 | L Frontal pole | --- | HC | HC– DID | 55 | -4, 68, -14 | **2.68 (0.009)\*** | -0.45 (0.657) | 0.72 (0.475) |
| 23 | R Inferior Occipital gyrus | --- | HC | HC-DID | 65 | 46, -74, -2 | **3.39 (0.001)\*** | 1.02 (0.311) | 0.16 (0.875) |
| 24 | L/R Mid cingulate | cSN | HC | HC– DID | 75 | 2, -12, 42 | **2.97 (0.004)\*** | -0.49 (0.629) | 1.14 (0.259) |
| 25 | L Precuneus, superior occipital gyrus | cSN | HC | HC– DID | 77 | -16, -64, 34 | **2.05 (0.043)\*** | 1.30 (0.196) | -1.55 (0.125) |
| 26 | L Frontal Opercular | cSN | HC | HC– DID | 91 | -36, 32, -2 | 0.93 (0.357) | -0.91 (0.364) | -0.17 (0.865) |
| 27 | L Middle Temporal Gyrus | tDN | HC | HC– DID | 457 | -48, -66, 8 | **6.38 (0.000)\*** | -1.35 (0.182) | **3.39 (0.001)\*** |
| 28 | L dmPFC | --- | HC | HC– DID | 485 | -8, 54, 10 | **2.97 (0.004)\*** | -0.13 (0.894) | 0.10 (0.924) |
| 29 | R Angular gyrus | rCEN | DID | DID– HC | 16 | 32, -66, 48 | -0.90 (0.372) | 0.85 (0.399) | 0.09 (0.933) |
| 30 | R vIPFC, Frontal pole | rCEN | DID | DID– HC | 16 | 28, 52, 0 | -1.61 (0.112) | -0.76 (0.449) | 0.95 (0.347) |
| 31 | R dIPFC, Middle frontal gyrus | rCEN | DID | DID– HC | 686 | 30, 34,42 | -1.35 (0.182) | 0.66 (0.510) | 0.47 (0.642) |
| 32 | R dmPFC | rCEN | + | Dep/Der | 388 | 6, 42,34 | 0.62 (0.537) | -0.55 (0.585) | 1.97 (0.053) |
| 33 | R IOFC, Frontal pole | rCEN | + | Dep/Der | 459 | 30, 52, 0 | -0.50 (0.616) | -1.01 (0.317) | 1.83 (0.071) |
| 34 | R Angular gyrus, Inferior Parietal Lobule | rCEN | + | Dep/Der | 700 | 32, -66, 48 | 0.13 (0.894) | 0.48 (0.630) | 1.27 (0.208) |
| 35 | R dIPFC, Inferior, Middle, Superior frontal gyrus | rCEN | + | Dep/Der | 1837 | 32, 32, 40 | -0.21 (0.835) | 0.51 (0.611) | 1.30 (0.197) |
| 36 | L dPCC | --- | - | Dep/Der | 13 | -6, -50, 24 | -1.20 (0.232) | -0.96 (0.341) | -1.32 (0.192) |
| 37 | L Cuneus | --- | - | Dep/Der | 22 | -6,-80, 34 | 0.90 (0.372) | -1.95 (0.055) | 0.44 (0.659) |
| 38 | L dIPFC, Superior Frontal gyrus | --- | - | Dep/Der | 25 | -14, 46, 42 | 1.49 (0.140) | -1.70 (0.093) | 0.70 (0.484) |
| 39 | L Frontal pole/amPFC | --- | - | Dep/Der | 35 | -4, 68, -14 | **2.58 (0.012)\*** | -0.69 (0.491) | 0.87 (0.385) |
| 40 | L Middle Temporal Gyrus | tDN | - | Dep/Der | 43 | -48, -64, 10 | **3.99 (0.000)\*** | **-2.76 (0.007)\*** | **3.23 (0.002)\*** |
| 41 | L Precuneus, Superior Occipital Gyrus | cSN, tDN | - | Dep/Der | 49 | -16, -70, 28 | 1.40 (0.165) | 0.74 (0.464) | -1.38 (0.173) |
| 42 | L Frontal Opercular | cSN | - | Dep/Der | 60 | -40, 28, -2 | 0.72 (0.474) | -1.35 (0.181) | 0.002 (0.999) |
| 43 | L dmPFC | --- | - | Dep/Der | 348 | -6, 58, 8 | **2.40 (0.019)\*** | -0.45 (0.652) | -0.10 (0.923) |
| 44 | R Angular Gyrus, Inferior Parietal Lobule | rCEN | + | PDI | 1125 | 32, -66, 48 | 0.40 (0.692) | 0.06 (0.950) | 1.60 (0.113) |
| 45 | R IOFC, Middle frontal gyrus, Superior Frontal gyrus | rCEN | + | PDI | 4074 | 50,38, 24 | 0.26 (0.795) | -0.42 (0.673) | **2.21 (0.030)\*** |
| 46 | L IOFC | --- | - | PDI | 9 | -42, 36, -12 | 0.07 (0.943) | -0.89 (0.374) | 0.27 (0.791) |
| 47 | L Postcentral gyrus | --- | - | PDI | 11 | -50, -20, 24 | 1.27 (0.207) | 0.00 (1.000) | -0.90 (0.369) |
| 48 | L IOFC | cSN | - | PDI | 13 | -50, 24, -10 | -0.09 (0.926) | -0.24 (0.808) | -0.49 (0.624) |
| 49 | L Cuneus | --- | - | PDI | 26 | -6, -80, 34 | 0.94 (0.352) | -1.86 (0.066) | 0.45 (0.658) |
| 50 | L Superior Temporal Gyrus | --- | - | PDI | 32 | -60, -52, 22 | 1.48 (0.142) | 0.26 (0.793) | -1.24 (0.220) |
| 51 | L Temporal-parietal-occipital junction | tDN | - | PDI | 36 | -36, -66, 24 | **3.39 (0.001)\*** | **2.23 (0.028)\*** | -0.84 (0.406) |
| 52 | L Inferior frontal gyrus | --- | - | PDI | 36 | -50, 20, 22 | 1.73 (0.088) | -0.62 (0.539) | 0.44 (0.659) |
| 53 | L Middle Temporal | tDN | - | PDI | 46 | -48, -64, 10 | **3.82 (0.000)\*** | **-2.38 (0.021)\*** | **2.87 (0.005)\*** |
| 54 | L dACC | cSN | - | PDI | 61 | -8, 20, 38 | 1.06 (0.293) | 0.11 (0.915) | -0.86 (0.392) |
| 55 | L Frontal operculum | cSN | - | PDI | 157 | -36, 32, -2 | 0.69 (0.489) | -0.70 (0.489) | -0.39 (0.695) |
| 56 | L mOFC, vmPFC | tDN | - | PDI | 256 | -2, 66, 12 | **2.13 (0.036)\*** | 0.46 (0.645) | -0.32 (0.753) |
| 57 | L dPCC, Precuneus | tDN | - | PDI | 306 | -16, -70, 26 | 0.54 (0.591) | 0.49 (0.626) | **-2.20 (0.030)\*** |
| 58 | L dmPFC | --- | - | PDI | 665 | -6, 54, 8 | **2.01 (0.047)\*** | -0.26 (0.798) | -0.45 (0.653) |

**Figure 2.**
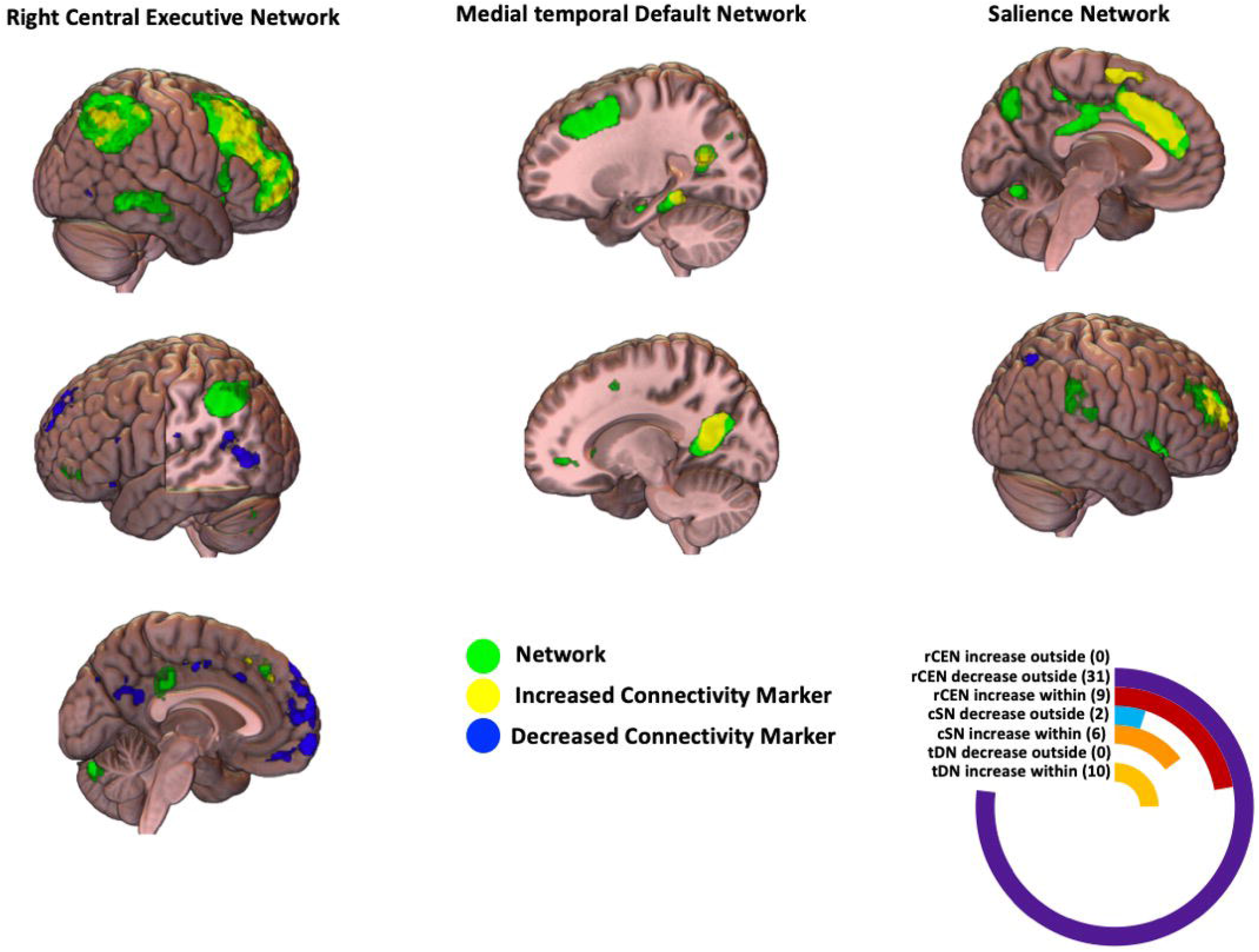
Triple Network Model of Pathological Dissociation. The Triple Network Model of Pathological Dissociation depicts biomarkers (brain regions) with functional connectivity to our core networks (right central executive, medial temporal default network, and salience network) that is associated with the full variance of each pathological dissociation variable (dissociative identity disorder diagnosis, depersonalization/derealization, and partially-dissociated intrusions). Green regions indicate the network of interest (right central executive, medial temporal default network, or salience network). Yellow indicates areas with increased connectivity between that region and the network of interest that is associated with pathological dissociation. Blue indicates regions with decreased connectivity between that region and the network of interest that is associated with pathological dissociation. The radial bar graph depicts the number of markers linked with pathological dissociation in each network associated with increased or decreased connectivity either within or outside the network of interest. Images made with MRIcroGL (https://www.nitrc.org/plugins/mwiki/index.php/mricron:MainPage). rCEN, right central executive network; cSN, cingulo-opercular salience network; tDN, medial temporal default network.

All three dissociation subtypes uniquely contributed to the altered connectivity of the rCEN (**Fig. 3, Table 2**). DID was associated with increased functional connectivity between rCEN and regions in tDN (cluster #40, 51, 53, 56), and with regions outside our three core networks (cluster #39, 43, 58). DID was also uniquely associated with decreased functional connectivity between rCEN and regions in tDN (cluster #27), cSN (cluster #24, 25), and regions outside our three networks (cluster #20, 22, 23, 28). Greater partially-dissociated intrusions were associated with rCEN within-network hyperconnectivity concentrated in lateral orbitofrontal cortex, middle and superior frontal gyrus (cluster #45), increased connectivity between rCEN and regions in tDN (cluster #27, 40, 53) and decreased connectivity between rCEN and posterior cingulate cortex/precuneus, also in tDN (#57). Greater depersonalization/derealization was associated with decreased connectivity between middle temporal gyrus and rCEN (cluster #40, 53), and increased connectivity between temporal-parietal-occipital junction and rCEN (cluster #51).

**Figure 3.**
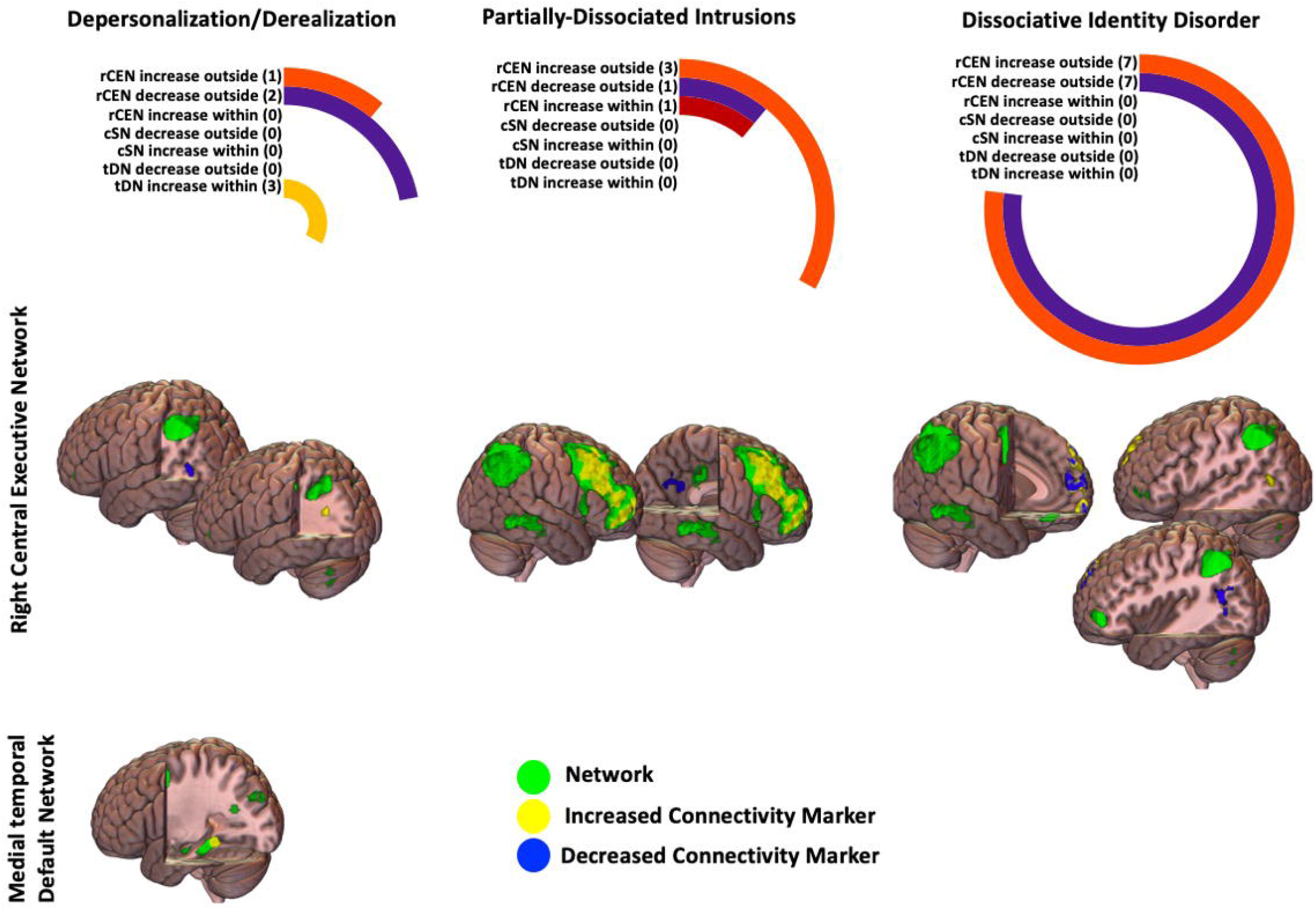
Unique Associations between Connectivity Biomarkers and Depersonalization/Derealization, Partially-Dissociated Intrusions and Dissociative Identity Disorder Diagnosis. Here we depict biomarkers (brain regions) with functional connectivity to our core networks (right central executive, medial temporal default network, and salience network) that is uniquely associated with each of the pathological dissociation variables (dissociative identity disorder diagnosis, depersonalization/derealization, and partially-dissociated intrusions). Green regions indicate the network of interest (right central executive, medial temporal default network). Only two of the three networks are shown because no markers with functional connectivity to salience network were uniquely associated with one of the dissociation variables. Yellow indicates regions with increased connectivity between that region and the network of interest uniquely associated with the dissociation variable. Blue indicates regions with decreased connectivity between that region and the network of interest with the dissociation variable. The radial bar graphs depict the number of markers linked with each type of dissociation in each network. The markers reflect either increased or decreased connectivity of regions within or outside the network of interest. Images made with MRIcroGL (https://www.nitrc.org/plugins/mwiki/index.php/mricron:MainPage). rCEN, right central executive network; cSN, salience network; tDN, medial temporal default network.

### Medial Temporal Default Network (tDN)

Ten clusters within tDN exhibited within-network hyperconnectivity related to pathological dissociation (**Fig. 2**). Only depersonalization/derealization showed unique associations with tDN connectivity, reflecting hyperconnectivity in parahippocampal gyrus (cluster #1, 3, 4; **Fig. 3**, **Table 2**).

### Cingulo-opercular Salience Network (cSN)

Eight clusters within cSN were linked to greater pathological dissociation in two ways (**Fig. 2, Table 2**): 1) within-network hyperconnectivity; 2) decreased connectivity between regions in rCEN with cSN. There were no significant unique contributions of dissociation subtypes.

## Discussion

To begin to build a large-scale functional network connectivity model of pathological dissociation and its subtypes, we leveraged the Triple Network model of psychopathology. We tested the connectivity of three core neurocognitive networks as it related to DID and the dimensional subtypes of depersonalization/derealization and partially-dissociated intrusions. Consistent with our hypotheses, after controlling for age, childhood maltreatment and PTSD symptom severity, the rCEN, tDN, and cSN were all impacted by pathological dissociation.

### Triple Network model of pathological dissociation

First, we examined alterations in functional connectivity related to pathological dissociation broadly defined as an association with DID diagnosis, depersonalization/derealization, and/or partially-dissociated intrusions. While each brain region was identified using a specific full variance model for each subtype of dissociation, the different subtypes are highly collinear. Consequently, the findings could be driven by shared variance between the subtypes. Therefore, we discuss results in this section as alterations due to “pathological dissociation,” not a particular subtype.

Overall, the rCEN was the most impacted by pathological dissociation; however, we found that all core networks implicated in the Triple Network model of psychopathology were impacted. Specifically, pathological dissociation was associated with hyperconnectivity within rCEN, tDN, and cSN. Dissociation was also linked to decreased connectivity between rCEN and other brain regions, including areas within DN and cSN that may facilitate communication among networks. Furthermore, greater dissociation was related to decreased connectivity between cSN and rCEN regions. Taken together, these alterations may be an adaptive or compensatory response to childhood trauma and are a likely source of executive functioning differences, self-alteration experiences, and altered interoceptive/autonomic experiences reported by individuals with dissociative symptoms (5,35,36).

### Depersonalization/Derealization

We found that depersonalization/derealization was uniquely associated with connectivity in two networks: the rCEN and tDN. First, depersonalization/derealization was related to decreased connectivity between rCEN and lateral middle temporal gyrus regions typically located in the DN and thought to facilitate retrieval of semantic/conceptual knowledge (14). This finding may reflect decreased communication between these networks. In contrast, rCEN had increased connectivity with the temporal-parietal-occipital junction typically located in DN. This region is involved in mentalization, that is, reflecting on the mental states of others (14). It is also implicated in out-of-body experiences (37). Increased communication between this region and CEN may, in part, underlie feelings of detachment, strangeness, or unreality with one’s body or environment.

Second, depersonalization/derealization was also associated with hyperconnectivity within tDN concentrated in parahippocampal gyrus. Parahippocampal gyrus is part of the medial temporal lobe memory system and has demonstrated connectivity with areas of the brain involved in vision (38). Parahippocampal gyrus supports memory formation and retrieval, in particular, for episodic and autobiographical memory, and the context of an event (38). Specifically, parahippocampal gyrus facilitates processing of spatial information essential for navigating one’s environment (38). Heightened communication within this region of DN may facilitate altered spatial and perceptual experiences associated with depersonalization/derealization.

### Partially-dissociated intrusions

We found that partially-dissociated intrusions were linked to rCEN hyperconnectivity concentrated in lateral prefrontal cortex. This network is often active during cognitively challenging working memory, problem solving, and decision-making tasks (9). This implies that greater partially-dissociated intrusions are related to heightened communication within CEN. This hyperconnectivity may also reduce the flexibility of the network to engage with other networks.

Second, partially-dissociated intrusions were associated with increased connectivity between DN regions (middle temporal gyrus) and rCEN. DN is often suppressed while CEN is engaged (9). However, here we see some synchronization of these two networks. Intriguingly, this matches the subjective experience of partially-dissociated intrusions as “recurrent, jarring, involuntary intrusions into executive functioning and sense of self” (29).

In contrast, rCEN had decreased connectivity with tDN regions: the dorsal posterior cingulate cortex (dPCC) and precuneus, which may reflect decreased communication between these networks. These regions are involved in self-generated thought (14). In particular, the dPCC may serve to regulate global brain dynamics – helping to balance internally vs. externally focused attention and the breadth of attentional focus (i.e., narrow vs. broad; (39)). Furthermore, recent theories speculate dPCC may facilitate fast shifts between different mental states (39).

### DID Diagnosis

The unique contributions of a DID diagnosis to altered connectivity were concentrated in the rCEN. Specifically, DID diagnosis was associated with a complex pattern of both increased and decreased connectivity between rCEN and regions distributed across tDN, cSN, and other networks. The dominant finding was one of rCEN hyperconnectivity with regions in tDN. DN is often suppressed while CEN is engaged (9), but in DID we instead saw some synchronization of these networks.

A pattern of decreased CEN connectivity with regions in cSN also emerged in DID. SN may facilitate shifts between CEN and DN (13). Decreased communication between rCEN and cSN could impact the appropriate engagement or disengagement of CEN and DN (9).

Overall, these findings support a plausible mechanism underlying executive functioning difficulties and differences in DID. For example, individuals with DID often report experiences of amnesia and partially-dissociated intrusions (5). Interestingly, there have also been some reports of preserved or even enhanced executive functioning for individuals with dissociative disorders or high levels of dissociation in which they out-perform control participants on executive functioning, working memory and spatial memory tasks that are not emotionally-provocative (35,40,41). It may be that some of the altered rCEN connectivity we identified could facilitate this enhanced executive functioning in certain contexts. Future work involving tasks that elicit CEN activity are needed to sort out when and how these alterations may facilitate enhanced vs. diminished executive functioning.

### Limitations

While we provide robust evidence for alterations in resting-state networks associated with pathological dissociation, future task-based analyses that directly measure self-generated thought, memory, and salience detection are needed to aid the interpretation of these findings. We also limited our analyses to rCEN, tDN, and cSN. Our findings suggest alterations between these three networks and other networks play a role in pathological dissociation, but we did not test this network-to-network connectivity directly.

### Clinical Implications and Significance

Finally, while gaps remain, this study contributes new data supporting the neurobiological basis of dissociative symptoms as a disruption of brain networks. Moreover, we have begun to develop a network-based brain connectivity “fingerprint” (23) specific to different types of dissociation. In the future, these neuromarkers could be used to stratify samples for randomized control trials, to monitor recovery, or to target directly with neuromodulatory techniques as a treatment intervention itself.

Given the complex and highly subjective nature of these conditions, neurobiological evidence is critical to ensuring that individuals who experience dissociation receive timely assessment and treatment, as with any serious neuropsychiatric condition.

## Data Availability

All data produced in the present study are available upon reasonable request to the authors

## Acknowledgments

The authors would like to thank the study participants and the hospital staff for their assistance and support. This research was supported by the Julia Kasparian Fund for Neuroscience Research (LAML, CP, MLK) and the National Institute of Mental Health K01 MH118467 (LAML), R21 MH112956 (MLK), and R01 MH119227 (MLK).

## Disclosures

LAML reports unpaid membership on the Scientific Committee for the International Society for the Study of Trauma and Dissociation (ISSTD), grant support from the National Institute of Mental Health (NIMH), K01 MH118467, and the Julia Kasparian Fund for Neuroscience Research. Dr. Lebois also reports spousal IP payments from Vanderbilt University for technology licensed to Acadia Pharmaceuticals unrelated to the present work. Dr. Ressler has performed scientific consultation for Bioxcel, Bionomics, Acer, Takeda, and Jazz Pharma; serves on Scientific Advisory Boards for Sage and the Brain Research Foundation, and he has received sponsored research support from Takeda, Brainsway and Alto Neuroscience. He receives research funding from the NIH. Dr. Kaufman reports unpaid membership on the Scientific Committee for the ISSTD and grant support from the NIMH, R21 MH112956, R01 MH119227. JTB has received consulting fees from Verily Life Sciences, as well as consulting fees and equity from Mindstrong Health, Inc., unrelated the present work. ISSTD and NIMH were not involved in the analysis or preparation of the manuscript. All other authors have nothing to report.

## Notes

### Funding Statement

This study was funded by the Julia Kasparian Fund for Neuroscience Research (LAML, CP, MLK) and the National Institute of Mental Health K01 MH118467 (LAML), R21 MH112956 (MLK), and R01 MH119227 (MLK).

### Author Declarations

IRB of Mass General Brigham gave ethical approval for this work.

