## Supplemental Materials for "Categorical and dimensional brain network-based models of trauma-related dissociative subtypes"

**Trauma-related dissociative subtypes characterized with resting-state functional connectivity**

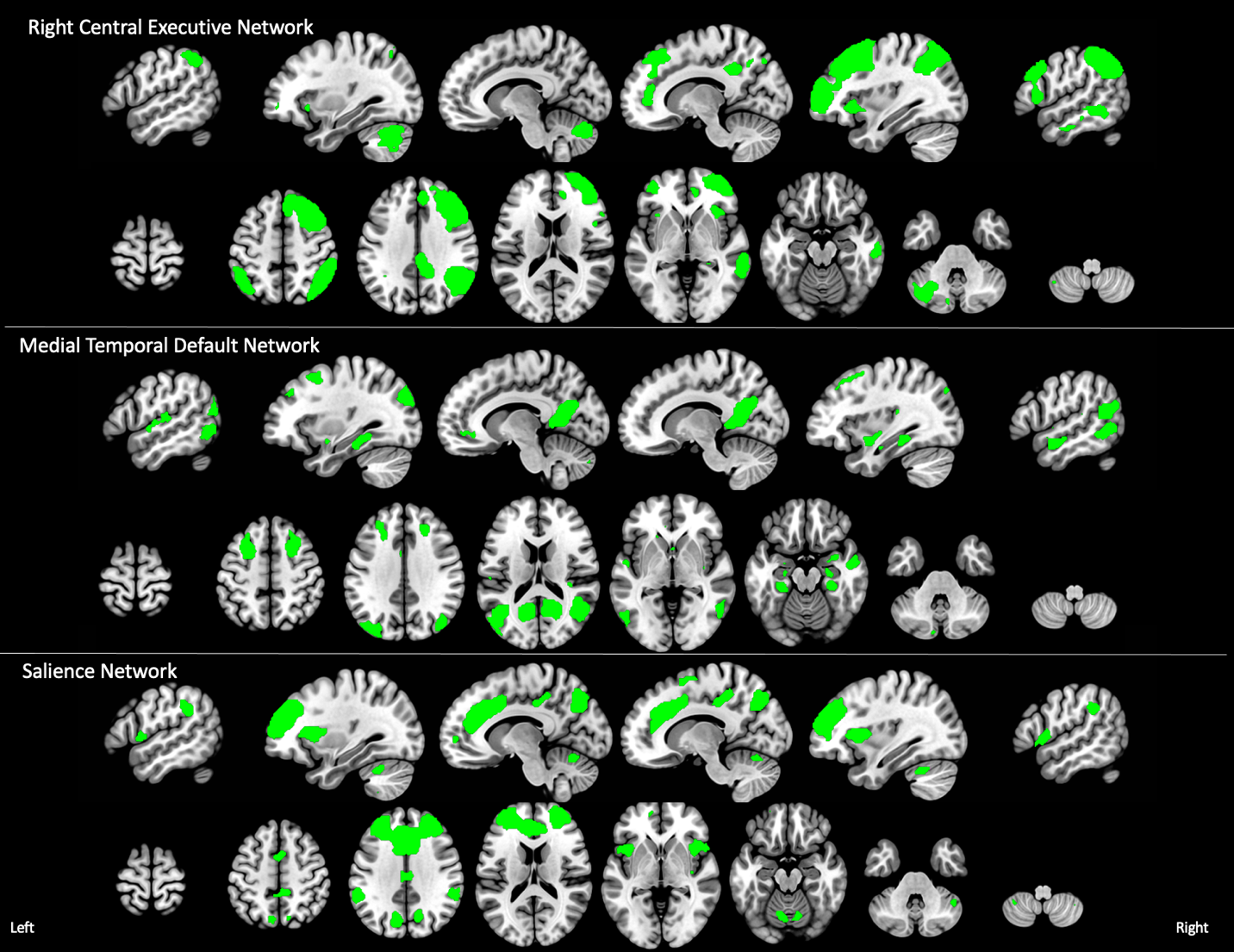

**Supplemental Figure 1**. The three resting-state networks of interest from the Triple Network Model. Slices go from left to right. x = -54, -32, -10, 12, 34, 54; z = -54, -36, -18, -2, 16, 34, 52, 70.

**Supplemental Methods**

**Diagnostic and Symptom Measures**. To measure partially-dissociated intrusions, we computed the average scores on the following Multidimensional Inventory of Dissociation (MID) subscales: child voices, voices/internal struggle, persecutory voices, speech insertion, thought insertion, made/intrusive emotions, made/intrusive impulses, made/intrusive actions, temporary loss of well-rehearsed skills and knowledge, disconcerting experiences of self-alteration, and self-puzzlement.

**MRI Procedures**

**Data Acquisition.** Blood oxygenation level dependent (BOLD) eyes-open resting-state fMRI data were collected using a 3T Trio scanner (Siemens, Inc), a 12-channel phased-array head coil and a 372 sec T2*-weighted echo-planar imaging sequence. The standard T2*-weighted echo-planar imaging had the following parameters: TR=3000 ms, TE=30 ms, flip angle=85°, 3x3x3mm voxels.  Participants were instructed to lie still with their eyes open and think about “nothing in particular” during the scan. A T1-weighted structural image was also collected for registration of fMRI data to standard space.

**Data Preprocessing.** Resting-state fMRI data were organized into BIDS format (1) reprocessed using *fMRIPrep* 20.0.1 (2,3).  fMRIPrep is based on Nipype 1.4.1 (4,5) and utilizes tools from both ANTS 2.2.0 (6) and FSL 5.0.11 (7).

For anatomical preprocessing, T1-weighted images were corrected for intensity non-uniformity with ANTs N4BiasFieldCorrection (8) then used as T1-weighted reference image throughout the workflow. The T1-weighted reference image for each participant was skull-stripped with a Nipype implementation of the ANTS-based antsBrainExtraction.sh workflow.  Brain tissue segmentation of cerebrospinal fluid, white-matter, and gray-matter was performed on the brain-extracted T1-weighted reference images using FSL’s FAST (FSL 5.0.11; (9)). Volume-based spatial normalization to the MNI152NLin6Asym standard space image was performed through nonlinear registration with antsRegistration.  This registration used brain-extracted versions of both the T1-weighted reference and the T1-weighted template.

For each subject’s BOLD fMRI data, the following preprocessing was performed: A reference volume and its skull-stripped version were generated using custom methodology in fMRIPrep. A deformation field to correct for susceptibility distortions was estimated based on fMRIPrep’s fieldmap-less approach (10). Registration of fMRI data to the T1-weighted references was performed with antsRegistration.  Based on the estimated susceptibility distortion, a corrected fMRI reference was calculated for a more accurate co-registration with the anatomical reference using FSL FLIRT (11) with boundary-based registration (12). Co-registration was done with nine degrees of freedom to account for distortions remaining in the BOLD reference.  Head-motion parameters with respect to the BOLD reference (transformation matrices, and six corresponding rotation and translation parameters) were estimated using FLIRT (13).  BOLD runs were slice-time corrected using 3dTshift from AFNI (14). The BOLD time-series were resampled onto their original, native space by applying a single, composite transformation to correct for head-motion and susceptibility distortions.  Pre-processed fMRI time-series were resampled into MNI152NLin6Asym standard space.

**Data Quality Assurance.** Several confound time-series were also calculated from the preprocessed native space BOLD data that were used for quality assurance: Framewise displacement (FD), the root mean square of the temporal change of the fMRI voxel-wise signal at each time point (DVARS), and three region-wise global signals.  Using Nipype, FD and DVARS were calculated for each functional run (15).  Motion outliers were frames that exceeded a threshold of 0.5 mm framewise displacement or 1.5 standardized DVARS.  Participants were excluded for excessive head motion (>18% of trials motion outliers).

We also conducted hand editing of the ICA-AROMA components, followed by denoising of the refined labeled motion components from each participants fMRI data.  Specifically, ICA-AROMA is optimized to detect subject motion rather than other artifacts.  Consequently, during our visual inspection of the ICA components used for data denoising, we consistently identified components mainly driven by multiband artifacts that were not classified as noise by ICA-AROMA.  Author PK visually inspected components for each subject and reclassified those that were not identified by ICA-AROMA.  Non-aggressive denoising was then redone using the fslregfilt command with the new set of classification labels.

The first four non-steady state volumes were removed and temporal filtering was then applied (highpass, 150 sec).  We then multiplied the fMRI data with an MNI brain mask to exclude non-brain regions.  These masked data were used for further analyses.

**Estimating Resting-State Networks.** Parcellation into resting-state networks was done using a multivariate data-driven method for group Independent Component Analysis (GICA; (16)).  GICA identifies sets of independent components by separating signals of interest, that is, resting-state networks, from physiological noise and artifacts that are common across all subjects.  To determine the number of components estimated by GICA, we ran GICA with different model orders (30, 35, 40 and 45).  We then visually inspected each model to determine which number of components produced resting-state networks that most highly resembled previously reported resting state networks (17–19). Visual inspection is needed to ensure network correspondence as spatial cross correlation sometimes misidentifies corresponding networks (because at this model order, patterns are large-scale spatially distributed networks). Using this approach, the model with 40 components was selected as the parcellation scheme for further analysis.

We then implemented a dual regression approach with this parcellation to obtain subject-specific network maps (20). First, we extracted subject-specific time courses for the GICA components via multivariate spatial regression of the GICA spatial maps against each subject’s fMRI time-series.  Second, we regressed the set of network time courses against each participant’s fMRI data to estimate subject-specific spatial maps corresponding to each GICA component. These maps are comprised of regression coefficients at each voxel that denotes the functional connectivity of each voxel with the corresponding resting-state network represented by the GICA component.

**Categorical and Dimensional Connectivity Analysis.** Our aim was to identify functional connectivity associated with a DID diagnosis, depersonalization/derealization and partially-dissociated intrusion severity.  Moreover, we wished to identify connectivity that was unique to each type of dissociation.  However, these subtypes are highly collinear.  Evaluating each type in separate models could yield findings that are driven by shared variance due to correlations between the variables (e.g., the overlapping area in the main text **Fig. 1**). On the other hand, if all three predictors were evaluated within the same model, this would reduce sensitivity as the shared variance between the predictors would be ignored in the estimation.

To address these issues, we followed the novel two-step approach presented in (21).  First, we conducted separate linear models for each predictor (DID diagnosis, depersonalization/ derealization, partially-dissociated intrusions) and network (right CEN, tDN, SN) to identify brain regions, or clusters, with connectivity to each network that was associated with the predictor of interest.  The predictor of interest was regressed from the other two predictors, and then the other two predictors were replaced by their residuals in the model.  This enabled us to capture the “full variance” associated with each predictor of interest (main text **Fig. 1**).

Next, for those clusters identified in the first step, we characterized the unique contribution of DID diagnosis, depersonalization/derealization, and partially-dissociated intrusions to the connectivity of that region with the network of interest.  We did this by performing the opposite of step 1: after we regressed out the two predictors from the one of interest, we replaced the predictor of interest with its own residuals (main text **Fig. 1**).

We applied this two-step strategy to the set of subject-specific spatial maps for each of the three *a priori* networks, tDN, right CEN, SN, as dependent variables in general linear models.  These models implemented steps one and two for the following variables of interest: the four diagnostic subgroups (PTSD, PTSD dissociative subtype, DID, and healthy control) and two additional symptom scores (depersonalization/derealization, partially-dissociated intrusion symptoms). Note that the models assessing diagnosis included all four diagnostic subgroup predictors.  In addition, age, CTQ childhood maltreatment severity, and CAPS-5 PTSD symptom severity were entered as covariates of no interest in all models.  Every model was evaluated using FSL Randomize for non-parametric permutation testing (*n*=5000 permutations) with threshold-free cluster enhancement to control family-wise error (*p*<.05).
